# Efficient Maternal to Neonatal transfer of SARS-CoV-2 and BNT162b2 antibodies

**DOI:** 10.1101/2021.03.31.21254674

**Authors:** Ofer Beharier, Romina Plitman Mayo, Tal Raz, Kira Nahum Sacks, Letizia Schreiber, Yael Suissa-Cohen, Rony Chen, Rachel Gomez-Tolub, Eran Hadar, Rinat Gabbay-Benziv, Yuval Jaffe Moshkovich, Tal Biron-Shental, Gil Shechter-Maor, Sivan Farladansky-Gershnabel, Hen Yitzhak Sela, Hedi Benyamini Raischer, Nitzan Dana Sela, Debra Goldman-Wohl, Ziv Shulman, Ariel Many, Haim Barr, Simcha Yagel, Michal Neeman, Michal Kovo

## Abstract

**Background:** The significant risks posed to mothers and fetuses by COVID-19 in pregnancy have sparked a worldwide debate surrounding the pros and cons of antenatal SARS-CoV-2 inoculation, as we lack sufficient evidence regarding vaccine effectiveness in pregnant women and their offspring. We aimed to provide substantial evidence for the effect of BNT162b2 mRNA vaccine versus native infection on maternal humoral, as well as transplacentally acquired fetal immune response, potentially providing newborn protection.

**Methods:** A multicenter study where parturients presenting for delivery were recruited at 8 medical centers across Israel and assigned to three study groups: vaccinated (n=86); PCR confirmed SARS-CoV-2 infected during pregnancy (n=65), and unvaccinated non-infected controls (n=62). Maternal and fetal blood samples were collected from parturients prior to delivery and from the umbilical cord following delivery, respectively. Sera IgG and IgM titers were measured using Milliplex MAP SARS-CoV-2 Antigen Panel (for S1, S2, RBD and N).

**Results:** BNT162b2 mRNA vaccine elicits strong maternal humoral IgG response (Anti-S and RBD) that crosses the placenta barrier and approaches maternal titers in the fetus within 15 days following the first dose. Maternal to neonatal anti-COVID-19 antibodies ratio did not differ when comparing sensitization (vaccine vs. infection). IgG transfer rate was significantly lower for third-trimester as compared to second trimester infection. Lastly, fetal IgM response was detected in 5 neonates, all in the infected group.

**Conclusions:** Antenatal BNT162b2 mRNA vaccination induces a robust maternal humoral response that effectively transfers to the fetus, supporting the role of vaccination during pregnancy.

## INTRODUCTION

The worldwide pandemic of coronavirus disease 2019 (COVID-19) continues to spread, with substantial morbidity and mortality. To date, over 80,000 pregnant women have been infected in the U.S. alone, and the estimated global number of pregnant women infected with COVID-19 is likely to reach millions this year. Recent data demonstrated that pregnant women with COVID-19 infection are at increased risk for intensive care unit (ICU) admission, mechanical ventilation, and death, compared with properly matched non-pregnant women ^1-5 1 6 7 8 9^. Furthermore, COVID-19 illness increases the risk for pregnancy complications, such as preterm birth, pregnancy-induced hypertensive diseases, and thromboembolic diseases ^10^. Although accumulating data suggest that the risk for severe morbidity and mortality among infected pregnant women is low, the Center for Disease Control and Prevention (CDC ^11^) included pregnancy as a risk factor for severe COVID-19 illness; a statement that was also acknowledged by the American College of Obstetricians and Gynecologists (ACOG), the Society for Maternal-Fetal Medicine (SMFM), and other women’s health organizations ^12 10 13^.

An important aspect of the maternal response to COVID-19 infection is the rapid resolution of infection by neutralizing immune response and transfer of immunity to the newborn. The maternal immune system plays a unique role in pregnancy, since the newborn depends on the active transfer of maternal immunoglobulin G (IgG) across the placenta for its protection against pathogens ^14 15 16^. Recent data revealed decreased placental transfer of COVID-19 specific antibodies ^17^, secondary to altered glycosylation profile ^17 18^. Proper transfer of neutralizing antibodies may be critical during pregnancy, as a greater proportion of neonates and infants have severe or critical illness upon COVID-19 infection than older pediatric counterparts ^19 20^.

In an attempt to stop the COVID-19 pandemic spread, mass vaccination campaigns commenced worldwide. Randomized clinical trials reported efficacy of 94% ^21^ to 95% ^22^for mRNA-based vaccines; however, these studies excluded pregnant women. Nevertheless, following extensive discussion regarding the risk of COVID-19 during pregnancy, potential vaccine benefits and safety concerns, the CDC, the Israel Ministry of Health, and other health organizations advised that the COVID-19 vaccine should be offered to pregnant women. Accordingly, a vaccination campaign among pregnant women in Israel began in December 2020. Herein, our objective was to evaluate the maternal production and placental transfer of antibodies following vaccination with the BNT162b2 mRNA vaccine and natural SARS-CoV-2 infection during pregnancy. We analyzed BNT162b2 mRNA vaccine-induced IgG and IgM antibody concentrations in maternal and cord blood samples from 105 deliveries at eight medical centers in Israel, between January and March 2021. Furthermore, we compared these results to IgG and IgM antibody concentrations in maternal and cord blood samples from 74 deliveries of women with PCR confirmed SARS-CoV-2 infection, contracted during various stages of pregnancy, as well as to 62 non-infected unvaccinated matched pregnant controls.

## METHODS

The current study followed the Strengthening the Reporting of Observational Studies in Epidemiology (STROBE) reporting guideline. The study was approved by the institutional review boards of all participating medical centers and by the Weizmann Institute of Science. All research participants provided written informed consent prior to enrollment.

### Study Design

Pregnant women admitted for delivery at eight medical centers in Israel (Hadassah Mt. Scopus, Wolfson, HaEmek, Hillel Yafe, Rabin, Shaare Zedek, Meir, and Sourasky Medical Centers) were approached for enrollment in a biorepository study, starting in April 2020. Eligibility criteria included an age of 18 years or older, and a willingness to participate and provide informed consent. Pregnant women with active maternal COVID-19 disease at delivery were excluded from this study. Eligible patients were identified by dedicated study clinicians (obstetrician, nurse midwife) present on the Labor and Delivery units enrolled in the study. Gravidae who received the BNT162b2 mRNA vaccine during pregnancy were assigned to the vaccinated group; parturients with documented COVID-19 infection during pregnancy, confirmed by positive nasopharyngeal swab RT-PCR test, comprised the COVID-19 positive group. Unvaccinated parturients were matched to the vaccinated group participants based on clinical parameters (Table 1) and were assigned to a control group.

**Table 1.** Clinical parameters of women included in the study

| **Parameter\*** | **Control group**<br>**n=66** | **Past SARS-CoV-2 group**<br>**n=74** | **Vaccinated group**<br>**n=92** |
| --- | --- | --- | --- |
| Maternal age, mean±SD, y | 31.6±5.8 | 28.8±5.8* | 31.7±5.8 |
| Gestational age, mean±SD, wks | 39.2±1.4 | 39±1.6 | 39.3±1.3 |
| Preterm delivery (<37), No. (%) | 5 (7.6) | 8 (10.8) | 4 (4.3) |
| Pregravid BMI (kg/m<sup>2</sup>), mean±SD | 25.7±6.5 | 26.4±9.2 | 24.2±5.2 |
| Gravidity, median (IQR) | 3 (2.5) | 3 (2) | 3 (2) |
| Parity, median (IQR) | 2 (2) | 1 (3) | 1 (2) |
| Maternal comorbidities, No. (%) |  |  |  |
| Hypertensive disorders | 1(1.5) | 1 (1.4) | 1(1.1) |
| Diabetes or gestational diabetes | 9 (13.6) | 4 (5.4) | 8 (8.7) |
| Asthma | 1 (1.5) | 2 (2.7) | 2 (2.2) |
| Thyroid disease | 4 (6.1) | 1 (1.3) | 8 (8.6) |
| Smoker | 4 (6.6) | 1 (1.4) | 6 (6.5) |
| Infant sex, No. (%) |  |  |  |
| Male | 31 (47.7) | 39 (52) | 45 (49.5) |
| Female | 34 (52.3) | 36 (48) | 46 (50.5) |
| Birthweight, mean±SD, gr | 3239.4±395.7 | 3324.1±536.2 | 3281.8±420.2 |
| NICU, No. (%) | 1 (1.6) | 2 (2.7) | 4 (4.3) |
| PCR-Positivity GA, mean±SD, wks | ----- | 28.1±8.3 | ----- |
| 1st Vaccine Dose GA, mean±SD, wks | ----- | ----- | 34.5±7.5 |

\*Clinical parameters did not differ among the groups, except for maternal age, which was significantly lower in the SARS-CoV-2 group, as compared to the other two groups (Kruskal-Wallis One-Way ANOVA; P=0.0011)

### Samples Collection and handling

Maternal and fetal blood samples were collected from the enrolled patients prior to delivery, and from the umbilical cord following delivery, respectively. The umbilical cord was wiped clean, and blood was drawn from the vein. Blood samples were centrifuged at 1000g for 10 minutes at room temperature, and serum samples were aliquoted into dedicated pre-coded tubes and stored at - 80°C until analyses.

### Quantification of Anti-COVID-19 Antibodies

Serum samples were thawed, heat-inactivated at 56°C for 30 minutes, and transferred to bar-coded 96-well plates for analysis. Serum IgG and IgM were detected using Milliplex MAP SARS-CoV-2 Antigen Panel 1 IgG (HC12SERG-85K) and IgM (HC19SERM1-85K). Reagents were prepared according to manufacturer instructions and dispensed to 96-well source plates (Greiner 651201, Sigma-Aldrich). Serum samples were diluted 1:100 in assay buffer and added to antigen-immobilized Milliplex beads in 96-well plates using a Bravo liquid handler (Agilent). Plates were covered, shook for two hours at room temperature, and washed three times with wash buffer, using a manual magnet and multidrop combi dispenser (Thermo). Anti-IgG-PE or Anti-IgM-PE conjugate was added, and the samples were incubated (90 minutes with shaking) and washed. Sheath fluid was added to the samples, and net fluorescent intensity (MFI) signals were detected on a Luminex MAGPIX reader.

### Statistical Analyses

Statistical analyses were performed using Statistix 8 software (Analytical Software, Tallahassee, FL USA) and Prism 5.01 (GraphPad Software; San-Diego, CA, USA). IgG and IgM antibody (S1, S2, RBD, N) concentrations were log10-transformed for analyses. Correlations between fetal and maternal Ab were analyzed by Linear Regression test. Comparisons of antibody concentrations among groups, as well as continuous parameters (e.g., clinical data), were analyzed by Kruskal– Wallis one-way ANOVA test, following by Dunn’s all-pairwise comparisons test; or alternatively, by Wilcoxon Rank Sum Test (if only two groups were compared). Comparisons between maternal and fetal concentrations within each group were analyzed by Paired t-test. Pearson Chi-square analysis was used to compare proportional data. All statistical testes were based on two-tailed hypotheses. Differences were considered significant at P < 0.05.

## RESULTS

### Participant characteristics

The cohort consisted of 1094 participants from eight hospitals across Israel. Samples were obtained following childbirth and stratified into three groups: 105 vaccine recipients, 94 un-vaccinated participants with past SARS-CoV-2 positive RT-PCR results, and 895 unvaccinated without prior documentation for infection (Table 1, Figure S1). Matched maternal cordblood serology results were obtained for 213 dyads; n=65 with past RT-PCR positive results, n=86 vaccinated recipient (3 of which were also PCR-positive during pregnancy), and n=62 non-infected unvaccinated; 55 enrolled participants did not have matched cord blood serological results for analysis and were therefore excluded from the analysis. Among the 895 non-infected unvaccinated participants, 66 were selected (based on clinical parameters) as matched comparison group and four were later excluded due to lack of matched maternal cordblood serology results. Participant demographic and clinical characteristics and outcomes are provided by study groups in Table 1. Clinical parameters did not differ among the groups, except for maternal age, which was significantly lower in the SARS-CoV-2 group, as compared to the other two groups (Kruskal-Wallis One-Way ANOVA; P=0.0011). Fourteen placental tissue samples were microscopically examined by a single experienced pathologist. The rate of malperfusion lesions was similar in the examined placental tissue of all groups (Supplementary).

### Maternal and fetal serological response to SARS-Cov2 infection

Based on serology analyses at delivery, transmission rates of IgG to S1, S2, RBD and N antigens were significantly higher in participants who were PCR positive to SARS-CoV2 prior to gestational week 30 (n=25), as compared to gestational week >30 (n=21) (Wilcoxon Rank Sum Test. S1, P=0.0013; S2, P=0.0231; RBD, P=0.0010; N, P= 0.0003). Maternal to fetal IgG transfer ratio values were consistently below 1 for gestational age >30 weeks, and were significantly elevated for earlier infections (Pearson’s Chi-Square, P<0.0001; Figure 1C).

**Figure 1.**
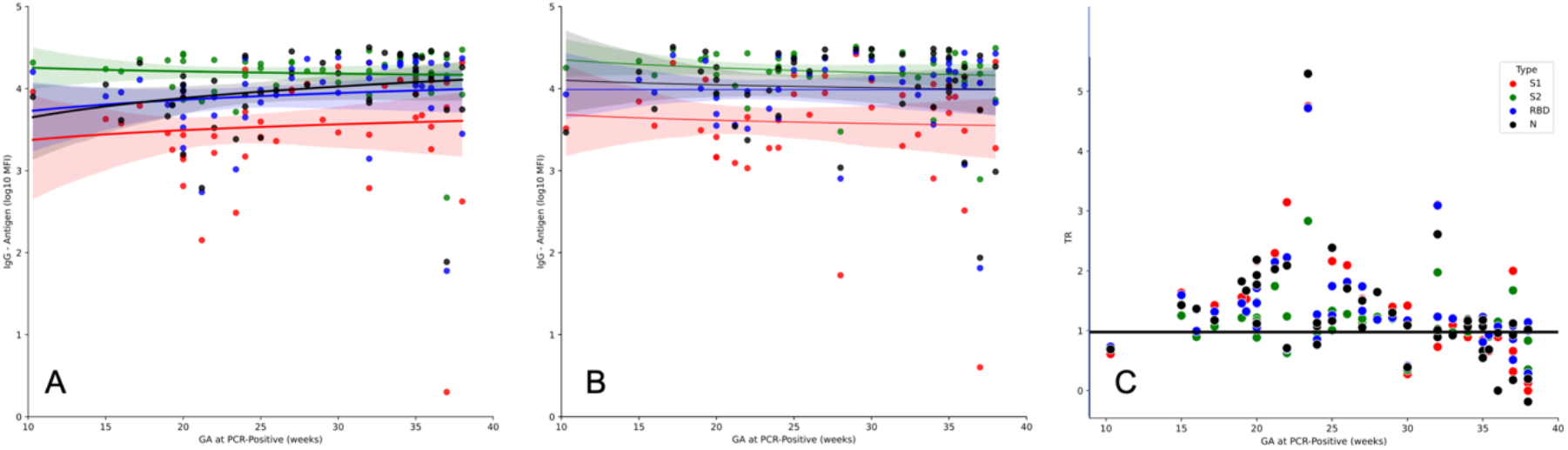
Robust maternal and fetal seropositivity is maintained after recovery from 2^nd^ trimester SARS-Co2 infection. Timeline of maternal and fetal seropositivity (A,B) and transfer ratio (C) for SARS-Cov-2 antigens (S1, S2, RBD and N) at the time of delivery, for patients (n=65) who recovered from infection with SARS-Cov-2 verified by RT-PCR during pregnancy. The data is plotted as a function of the gestational age (GA) at which positive RT-PCR was diagnosed. (A, B) High steady-state levels of maternal (A) and fetal (B) IgG levels for S1, S2, RBD and N are maintained until delivery, apart for a decline in late 3^rd^ trimester infections. (C) Transfer ratio values at delivery are low for 3^rd^ trimester infections, and significantly elevated for earlier, 2^nd^ trimester infections (P<001). S1, red; S2, green, RBD, blue; N, black. Shaded areas show the 95% confidence interval.

### Serological based re-clustering of the study groups

Based on the robust serological response maintained from mid-pregnancy PCR verified SARS-CoV2 infection until delivery, the multiplexed immune response was used for further clustering of the participants based on the reactivity to the N antigen. Multiplexed serological IgG and IgM response to S1, S2, RBD and N were tested in maternal and neonate sera (Figure S2). In particular, the response to N versus RBD separated the main groups and identified additional potentially infected participants of the vaccinated and control groups (Figure S3). Among 65 participants with past SARS-CoV2 RT-PCR positive test, the top 90% of the maternal IgG response for N was defined as seropositive (IgG N (MFI) > 1583; n=59). Within the control group, 9 participants (14%) were seropositive for N using the above threshold, which together with IgG seropositivity to the other COVID-19 antigens (S1, S2 and RBD), corresponds to a preexisting induced immunity due to infection. Similarly, 7 vaccinated participants (8%) were seropositive for N, of which 3 were also PCR-positive.

Notably, within the PCR positive group, 4 neonates were identified with robust IgM response to all SARS-CoV2 antigens, and an additional neonate showed partial response consistent with compromised placenta barrier, fetal exposure to viral antigens, or with vertical viral transmission. Clinical review of these cases showed that the mothers were diagnosed with mild SARS-CoV2 infection that spontaneously resolved weeks prior to childbirth. Three cases delivered at term, and one case gave birth at 35 weeks following preterm premature rupture of the fetal membranes. In all cases, both mother and newborn did not show any signs of illness after childbirth.

### Maternal and fetal serological response to BNT162b2 vaccine

The temporal dependence of the acute maternal response to SARS-CoV2 infection (days 1-45; Figure 2A) was compared to the response to the two-dose regime of BNT162b2; where the 1^st^ vaccine is administered on day 1 and the 2^nd^ dose on day 21 (Figure 2B). A gradual rise in IgG humoral response (Anti-S1, S2, RBD and N) was detected during the first 45 days after infection (Figure 2A). In the same period, vaccinated participants receiving the first BNT162b2 dose showed a rapid IgG response to S1, S2, RBD but not N, resulting in high titer values by day 15 after the first dose. A further rise in IgG was observed following the second dose (Figure 2B). The temporal dependence of fetal IgG for S1, S2 and RBD after vaccination trailed after the maternal IgG showing a significant response already by day 15. As expected, a further increase was observed following the second vaccination dose (Figure 2C). As illustrated in Supplementary Figure S4, at the time of delivery, maternal IgG for S1 and RBD were significantly higher in vaccinated women (P=0.0009, P=0.0045, respectively), while IgG for S2 and N were significantly higher in PCR-Positive women (P=0.0016, P<0.0001, respectively). Fetal IgG for S2 and N were significantly lower in bloodcords of vaccinated women (P<0.0001, P<0.0001, respectively), while fetal IgG for S1 and RBD did not differ from those of PCR-Positive women (P=0.7017, P=0.6887, respectively).

**Figure 2.**
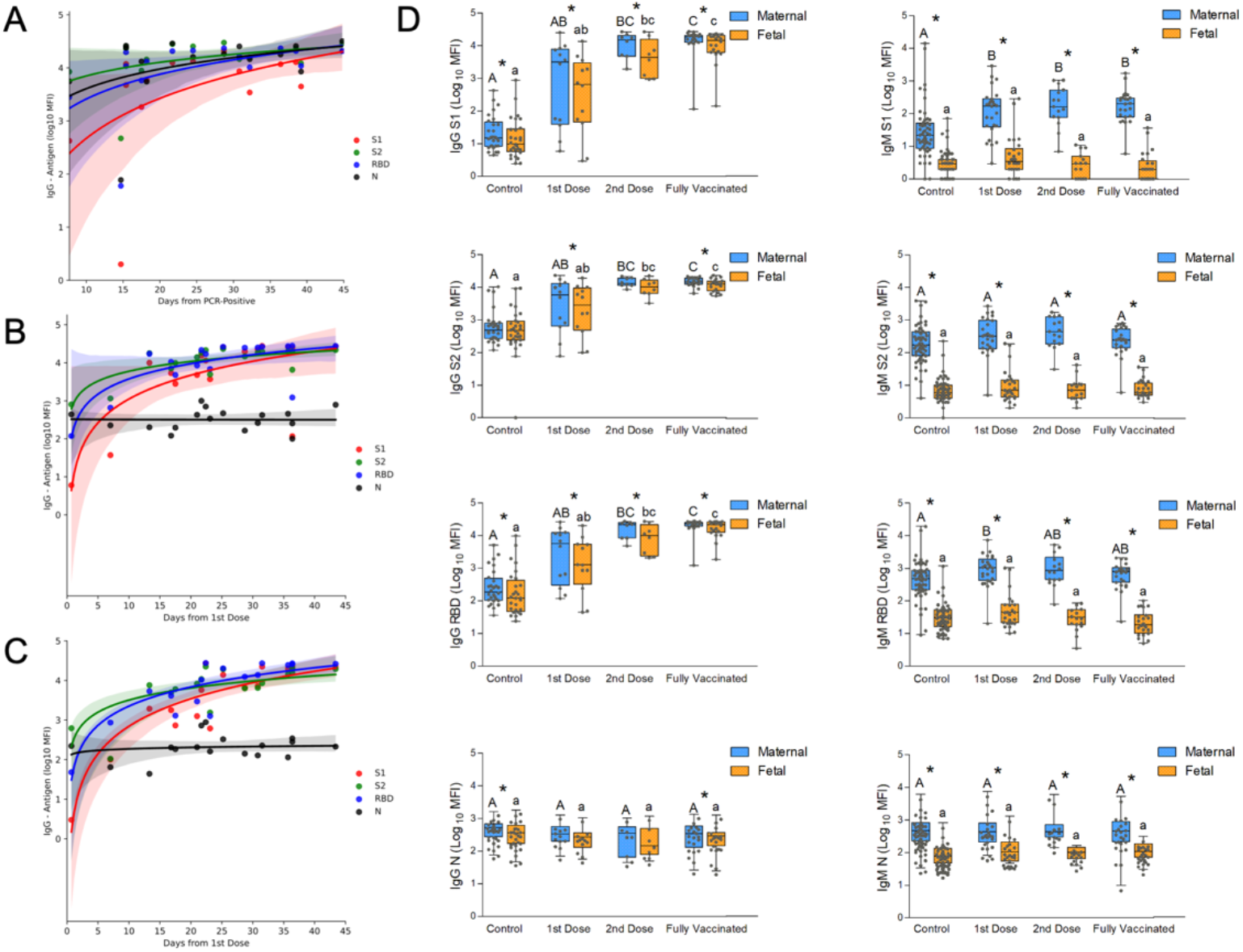
Temporal changes of the acute immune response to SARS-Cov2 infection and to vaccination in pregnancy. (A) Analysis of the change in maternal IgG during the first 50 days after positive RT-PCR, derived from the GA of positivity and the GA of delivery (see the full timeline data for immune response across pregnancy in Figure 1). (B) Analysis of maternal IgG response to BNT162b2 vaccination derived from the GA of the first vaccine and the GA of delivery. A second dose was administered on Day 21.. (C) Analysis of the temporal changes of fetal IgG following BNT162b2 vaccination derived from the GA of the first vaccine and the GA of delivery. A second dose was administered on Day 21; shaded areas in (A), (B) and (C) show the 95% confidence interval (D) Serological data of maternal-fetal pairs was derived from control, unvaccinated serologically negative (N-) mothers; as well as vaccinated mothers grouped for deliveries in the first 3 weeks after the 1^st^ vaccine; deliveries during the first week after the 2^nd^ vaccine; and fully vaccinated who delivered more than a week after the second vaccine. Left columns, IgG; right columns, IgM; from top to bottom, serological response to S1, S2, RBD, and N. Statistical significance: ^A,B,C^ above the blue bars indicate significant differences among the groups in maternal antibodies, while ^a,b,c^ above the orange bars indicate significant differences among the groups in fetal antibodies (Kruskall–Wallis one-way ANOVA test, following by Dunn’s All-Pairwise Comparisons Test); *indicate a significant difference between maternal and fetal antibodies within the same group (Paired t-test). Box and whiskers: Middle line= Median; Box= the 25^%^ and 75% (25th, and 75th percentiles); whiskers= min & max values (Table S2).

Paired maternal-neonate serological data was grouped for statistical analysis to control, unvaccinated mothers; as well as mothers who presented at delivery within the first 3 weeks after the 1^st^ vaccine; deliveries during the first week after the 2^nd^ vaccine; and fully vaccinated - deliveries more than 1 week after the second vaccine. Significant increase in maternal and fetal IgG (P<0.0001) and maternal IgM (P<0.05) to S1, S2 and RBD but not N were observed already after the first vaccination dose and persisted at later timepoints (Table S2). Fetal IgM response to BNT162b2 antigens (S1, S2, RBD) was negligible, consistent with no evidence for direct exposure of the fetus to vaccine-derived antigens (Figure 2D; Figure S2).

### Correlation of maternal-fetal IgG response to SARS-Cov2 infection and vaccination

Significant positive correlations were found between all fetal and maternal IgG antibodies (Figure 3), S1 (R^2^=0.9443; Adjusted R^2^= 0.9438; P<0.0001), S2 (R^2^= 0.9353; Adjusted R^2^= 0.9348; P<0.0001), RBD (R^2^= 0.9200; Adjusted R^2^= 0.9194; P<0.0001), and N (R^2^= 0.9366; Adjusted R^2^= 0.9361; P<0.0001). In all analyses, there were no differences between the correlation slopes of the SARS-Cov2 infected group vs. the vaccinated group in any type of antibodies (S1, P=0.2936; S2, P=0.4212; RBD, P=0.09702; N, P=0.7616), suggesting similar placental antibodies transfers following SARS-Cov2 infection and vaccination.

**Figure 3.**
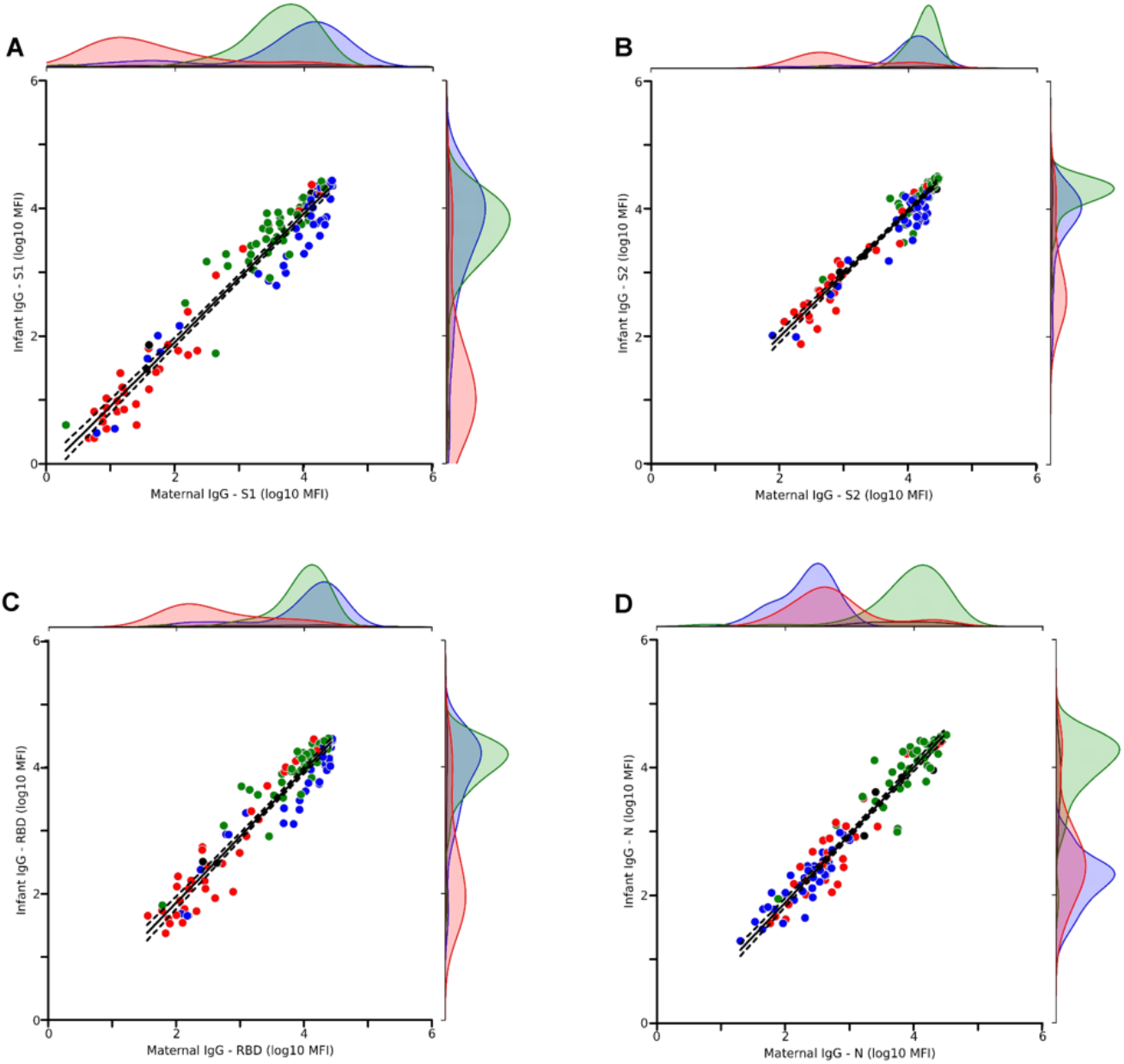
Maternal-fetal serological correlation of IgG for S1, S2, RBD and N. A subgroup of the Control group was identified as serologically positive for N as well as for S1, S2, and RBD (n=9 of 62). Similarly, a subgroup of the vaccinated was serologically identified as positive for N (n=7 of 86; marked in black). A significant maternal-fetal correlation was observed for all groups and all antigens. Correlations between fetal and maternal Ab (N, S1, S2, RBD) were analyzed by Linear Regression test (Supplementary Figure S3). Each dot represents data from a single patient; the linear regression line is marked in black, with its 95% CI (dotted lines). (A) R^2^=0.9443; Adjusted R^2^= 0.9438; P<0.0001. (B) R^2^= 0.9353; Adjusted R^2^= 0.9348; P<0.0001. (C) R^2^= 0.9200; Adjusted R^2^= 0.9194; P<0.0001. (D) R^2^= 0.9366; Adjusted R^2^= 0.9361; P<0.0001. Red, Control; Green, PCR positive; Blue, Vaccinated N-; Black, Vaccinated N+.

### Maternal to fetal IgG transfer ratio for S1, S2, RBD, and N

Maternal to fetal transfer ratio (TR), defined as fetal divided by maternal antibody levels, was derived for the PCR-positive group and for serologically positive and negative vaccinated groups (N^+^ and N^-^; Figure 4). Note that the TR for N in the N-group is not presented due to the low seropositivity. Significant differences were found for S1, S2 and RBD between the PCR-positive and vaccinated anti-N-groups (P<0.0002). The transfer ratios for all antibodies did not differ between the vaccinated anti-N^+^ and all the other groups (P=0.4577).

**Figure 4.**
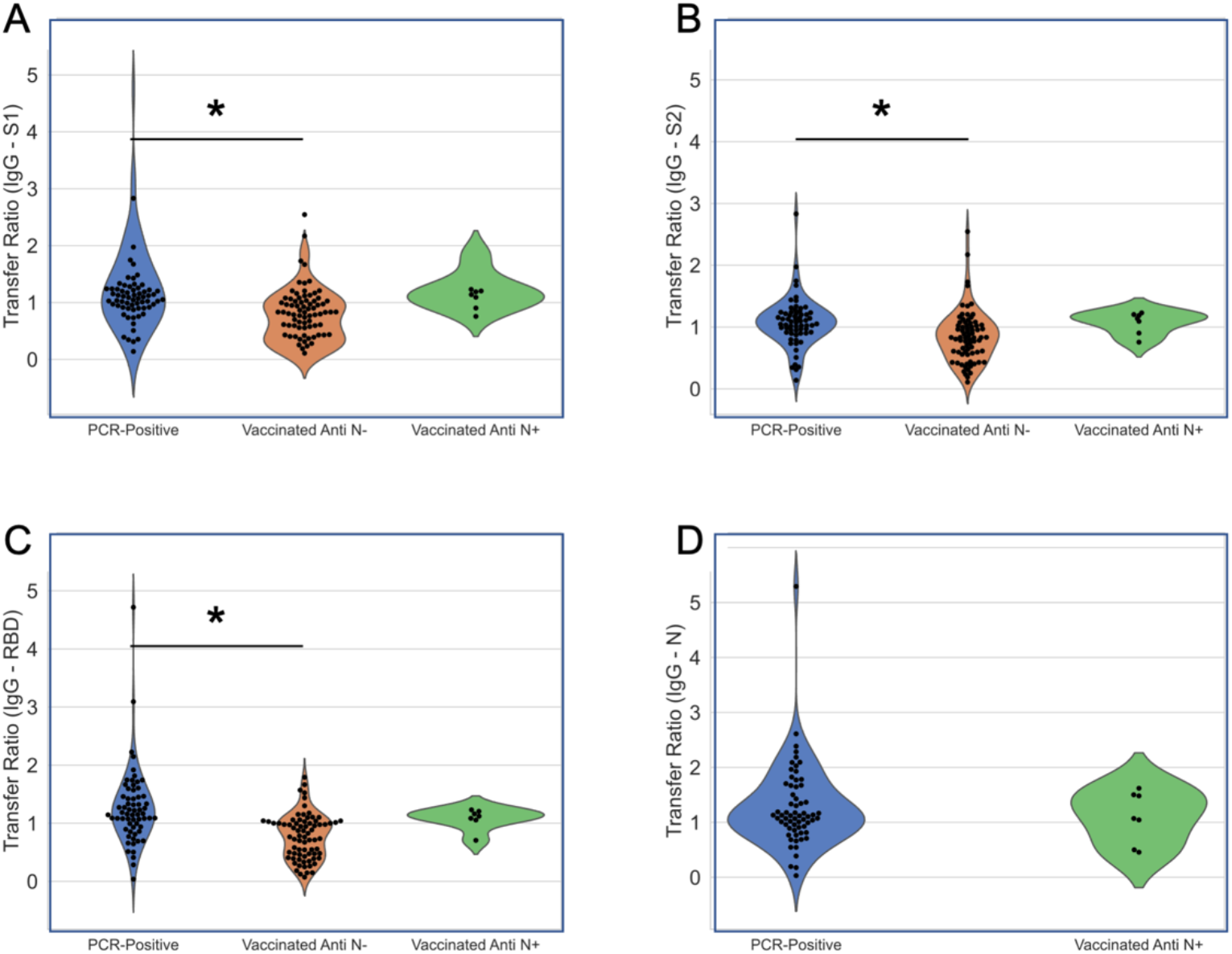
Maternal to fetal transfer ratio (TR) for the PCR positive recovered group and for serologically positive and negative vaccinated groups. Note that the TR for N in the N^-^ group is not presented due to the low seropositivity. Differences among the groups were analyzed by using Kruskall–Wallis one-way ANOVA test, following by Dunn’s All-Pairwise Comparisons Test. For S1, S2 and RBD, there was a significant difference between the PCR-positive vaccinated Anti N^-^ groups (P<0.0002). The transfer ratios did not differ between the vaccinated Anti N^+^ and all the other groups for all antigens (P=0.4577).

## DISCUSSION

Pregnant women and their neonates are considered vulnerable populations for COVID-19 infection, with significantly greater risks for morbidity and mortality, when compared to matched populations ^17^. Recent studies reported that among patients infected during the third trimester, the transfer of anti-SARS-CoV-2 antibodies to the fetus is significantly impaired ^17 18^. Indeed, our study confirmed the attenuated transfer ratio for infections late in pregnancy. However, with the availability of a large cohort of patients infected earlier in pregnancy (weeks 15-30) we were able to show for the first time that anti-COVID-19 antibodies generated in response to a second trimester infection remain high in maternal and cordblood until the time of delivery, in participants recovering from SARS-CoV-2 infection contracted months prior to childbirth.

Following reports of a decline in antibody titers months after infection, health organizations recently recommended vaccination following natural SARS-CoV-2 infection for boosting immunity ^23^. However, the significance and relevance of this policy during pregnancy is the subject of some debate and has not been supported by evidence. Based on the persistence of humoral immunity for infections contracted during the 2^nd^ trimester of pregnancy, we propose to consider vaccination of previously infected pregnant woman based on titer testing, unless boosting is warranted by emerging variants ^24^.

Unfortunately, pregnant women were excluded from previous clinical vaccine studies. However, the significant risks and pressing need for action led to a worldwide debate concerning SARS-CoV-2 inoculation during pregnancy, while data were still lacking. In the present study, we drew on the unprecedented vaccination campaign undertaken in Israel, which included pregnant women, and report on the robust humoral immune response following antenatal immunization with the mRNA vaccine. We found that the Pfizer-BioNTech COVID-19 mRNA vaccine elicits a rapid rise in IgG titers and effective transfer across the placenta, exceeding the TR observed in pregnant women with third trimester SARS-CoV-2 infection, as was previously described in non-pregnant populations, and in a small pilot study in pregnancy ^25^.

Importantly, maternal IgG humoral response to vaccination in non-infected patients readily transfers across the placenta to the fetus, leading to a significant, and potentially protective, anti-SARS-CoV-2 titer in the neonatal bloodstream, already two weeks following the first vaccine dose. Hence, our data delivers convincing proof for the potency of COVID-19 mRNA vaccines to induce robust humoral maternal and neonatal immunity during pregnancy. In addition to transplacental acquired humoral defense, other investigators have recently shown that vaccine response included the transfer of both spike-specific IgG and IgA antibodies into the maternal breastmilk, potentially building another line of defense for breastfed infants ^25^. Accordingly, antenatal immunization will potentially provide adequate maternal and neonatal protection at highly vulnerable life stages. Nevertheless, sound evidence regarding safety is still needed and should be addressed in future studies.

Utilizing multiplexed serology, we were able to distinguish between viral and vaccination induced immunity and uncover clusters of asymptomatic, undiagnosed infections among the control and vaccinated groups. We describe 7 vaccinated patients that were found to have high levels of anti-N IgG, corresponding with previous un-diagnosed infection. We detected no significant changes in maternal or neonatal titers, when compared to the titers of vaccinated and recovered participants. In addition, among the 65 PCR positive deliveries, we found 5 fetuses (7%) who showed IgM reactivity to all or most viral antigens, consistent with placenta barrier defect, fetal exposure to viral antigens, or vertical viral transmission. In contrast, among the 86 vaccinated deliveries, we found no evidence for cases of fetal IgM response to any of the vaccine-induced antigens.

### Strengths and Limitations

The present study has several strengths and limitations. Its strengths include its multicenter design and patient accrual; our relatively large cohort size; and our diverse patient population. Its limitations include the bias in sample collection as most of the study recruitment occurred during the day, and therefore does not includes most of the emergency cases. However, the method of sample collection did not differ between the study groups and Medical Centers, thus minimizing the impact of this effect on our results. Second, since sample collection began long before the COVID-19 immunization campaign, the duration of sample collection differed somewhat between the groups, with an extended recruitment period for COVID-19 recovered cases. We found no differences in background or demographic parameters among the groups. Third, a history of COVID-19 infection during pregnancy was made by positive RT-PCR results during pregnancy with self-reporting of the time of PCR test. Stricter and more accurate symptom monitoring and repeated sampling during pregnancy may provide a higher resolution delineation of how the immune response develops and transfers, following COVID-19 infection. Fourth, patients presenting with RT-PCR positive within one week prior to delivery were excluded from this study due to safety concerns. Thus, future studies are needed in order to characterize the maternal humoral response to COVID-19 within days of infection.

## Conclusions

We show herein a robust maternal humoral immune response coupled to a rise in protective antibodies in the fetal circulation as early as 15 days after the first BNT162b2 mRNA vaccination. We further show that mid-pregnancy SARS-Cov2 infection results in prolonged maternal and fetal humoral immunity presented at delivery time.

## Data Availability

Data will be made accessible after peer reviewed publication.

## Acknowledgements

This work was supported by ISF KillCorona grant 3777/19 (to MN, MK, SY, AM); by FERRING COVID-19 Investigational Grant in Reproductive Medicine and Maternal Health (RMMH) (to SY) and by a research grant from the Fondazione Henry Krenter (to MN). We would like to thank the patients who made this research possible. We acknowledge the contribution in patient recruitment, sample preparation and discussions by:

Weizmann Institute: Gila Meir, Leonardo Solmesky, PhD., Nava Dekel, PhD.

Hadassah Medical Center: Adva Cahen Peretz, MD; Michal Lipschuetz, RN, MPH, MSc; Nadine Souri; Sarah M Cohen, MPH.

Wolfson Medical Center: Yasmin Farhadian MD, Hind Odeh BSc

HaEmek Medical Center: Shalva Fux BA, RN (MW)

Hillel Yaffe Medical Center Alina Wiener, MBA, Luchilla Zorzetti, MD

Shaare Zedek Medical Center: Itamar Glick, MD

Meir Medical Center: Avital Diamond, Yaara Hoffman

