## Supplemental material for "Efficient Maternal to Neonatal transfer of SARS-CoV-2 and BNT162b2 antibodies"

**Table of contents**

| | |
|---|---|
| Figures | Page 2-6 |
| Tables | Page 7-11 |

Supplementary Figures

Supplementary Figure S1. Patient selection flow chart

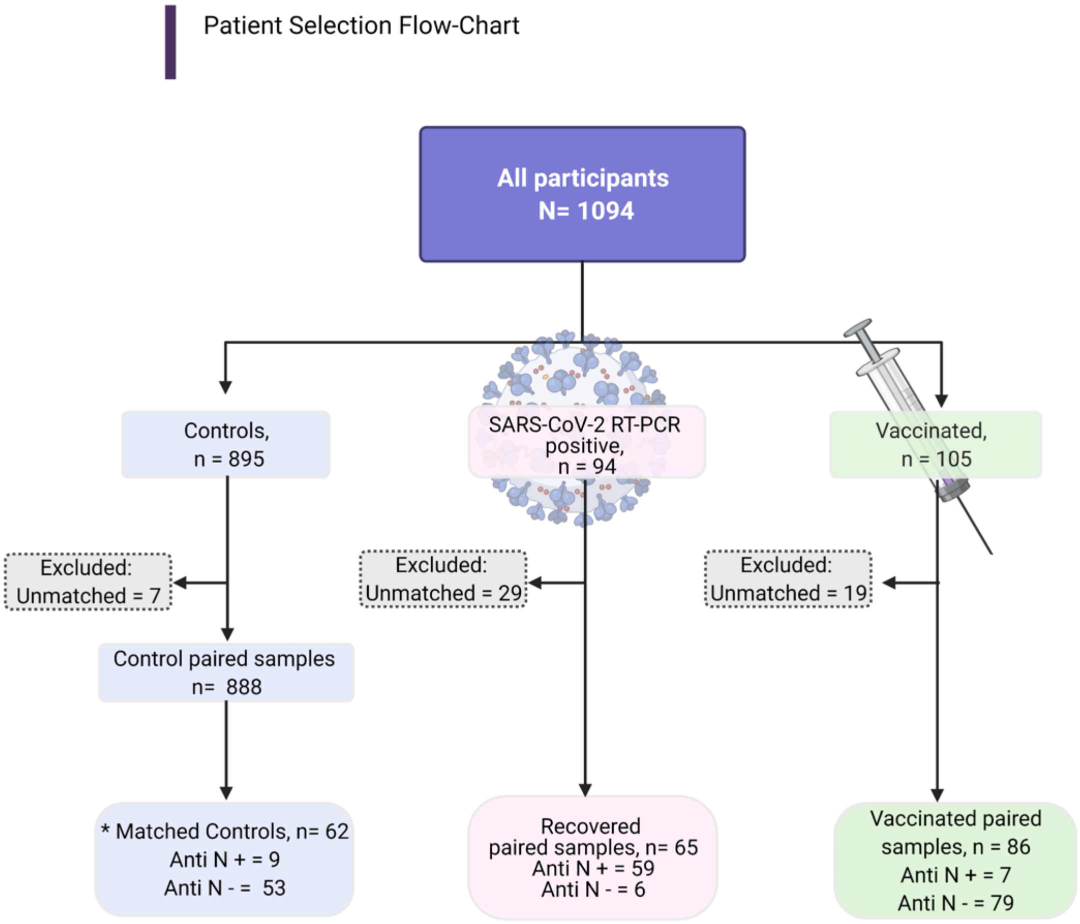

Figure S1 Patient selection flow chart. Patients were recruited from 8 Medical Centers in Israel and were all SARS-Cov-2 RT-PCR negative at delivery. Sero-positivity for nucleocapsid (N) was set at the level of the top 90% of the PCR-Positive, recovered group, and verified by positivity for S1, S2 and RBD (See supplementary Fig 2, 3).

Supplementary Figure S2

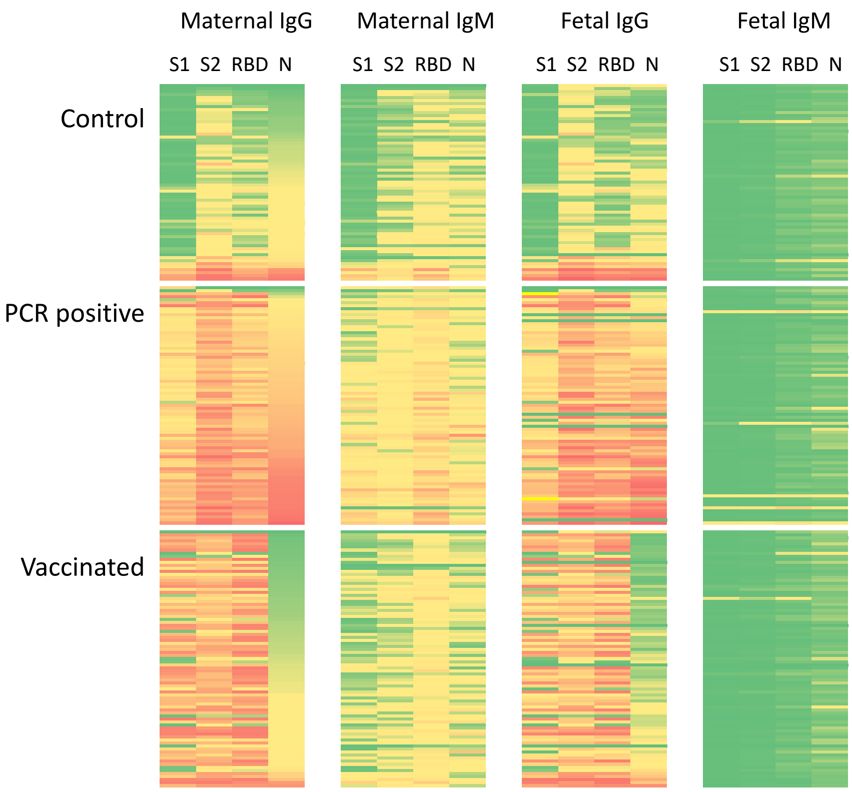

Figure S2 Serological heat map

The heat map was generated from the acquired IgG and IgM serological data segregated by the main recruitment groups (Control, Recovered, and Vaccinated). From left to right Maternal IgG (S1, S2, RBD, N), Maternal IgM (S1, S2, RBD, N), Fetal IgG (S1, S2, RBD, N) and Fetal IgM (S1, S2, RBD, N). Each row represents matched maternal-fetal data, ranked by the maternal IgG reactivity to N antigen within each group (Low, green; high, red). Note the serologically $N^+$ mothers (high N in red) within the control and the vaccinated groups.

Supplementary Figure S3

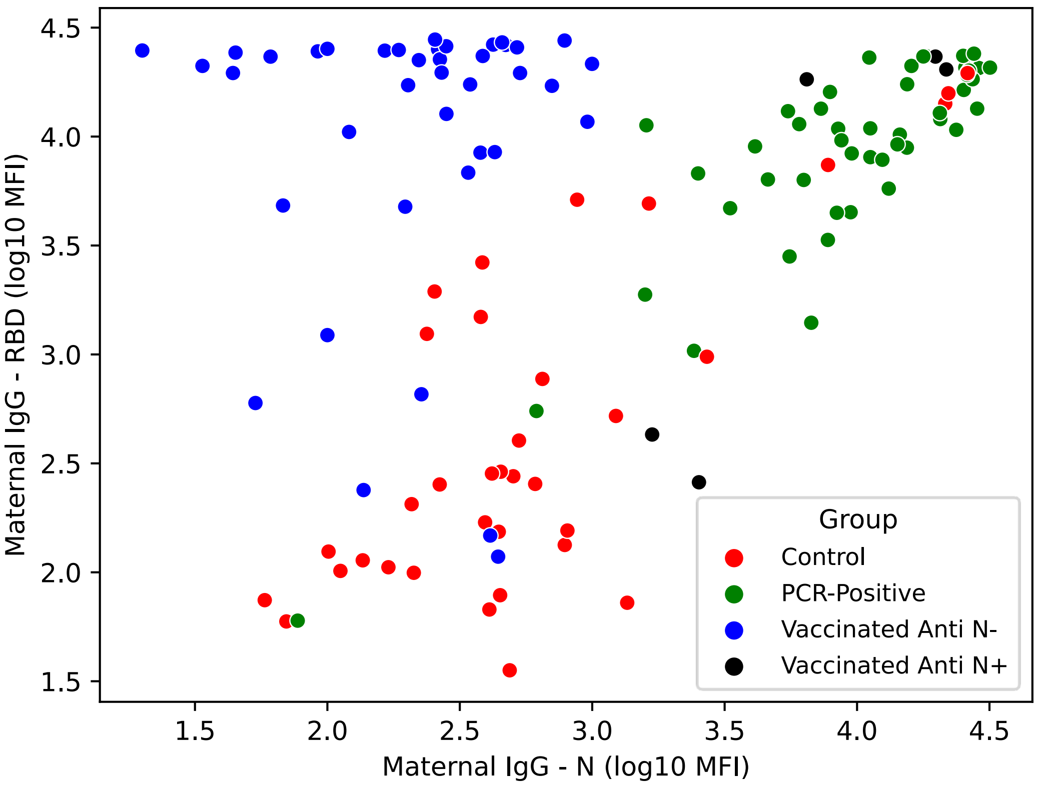

Figure S3 Cluster analysis of all study participants by maternal response to N and RBD.

Supplementary Figure S4

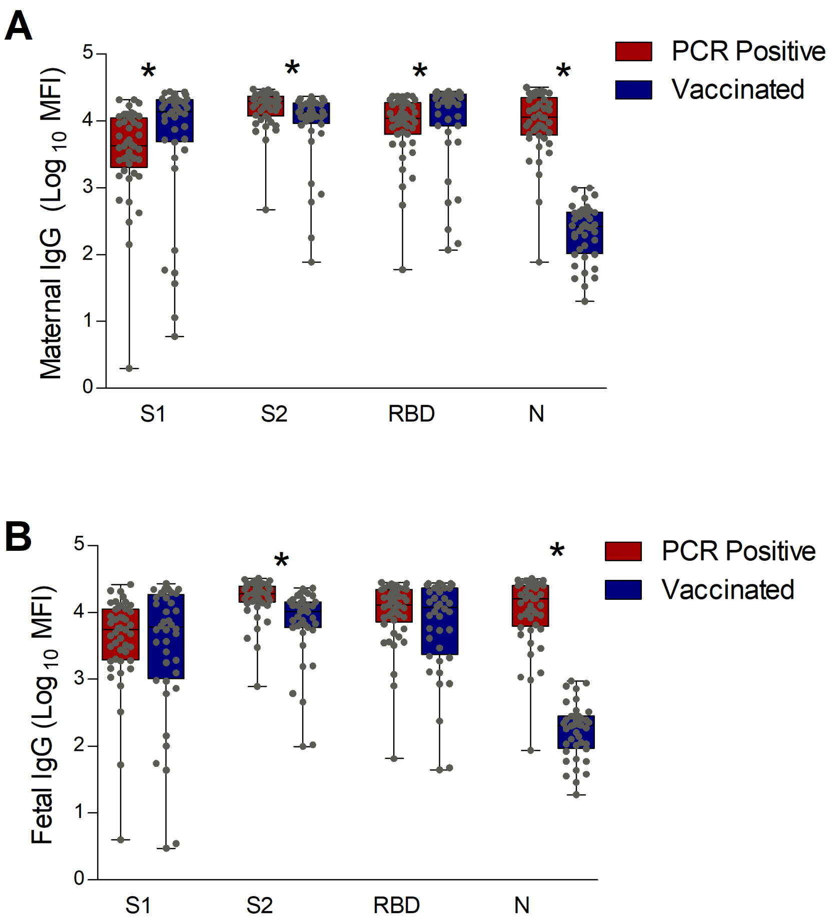

Figure S4. Maternal and fetal IgG for S1, S2 RBD and N following SARS-Co2 infection vs. vaccination.

Supplementary Figure S5 <u>Placental Samples Examination</u>

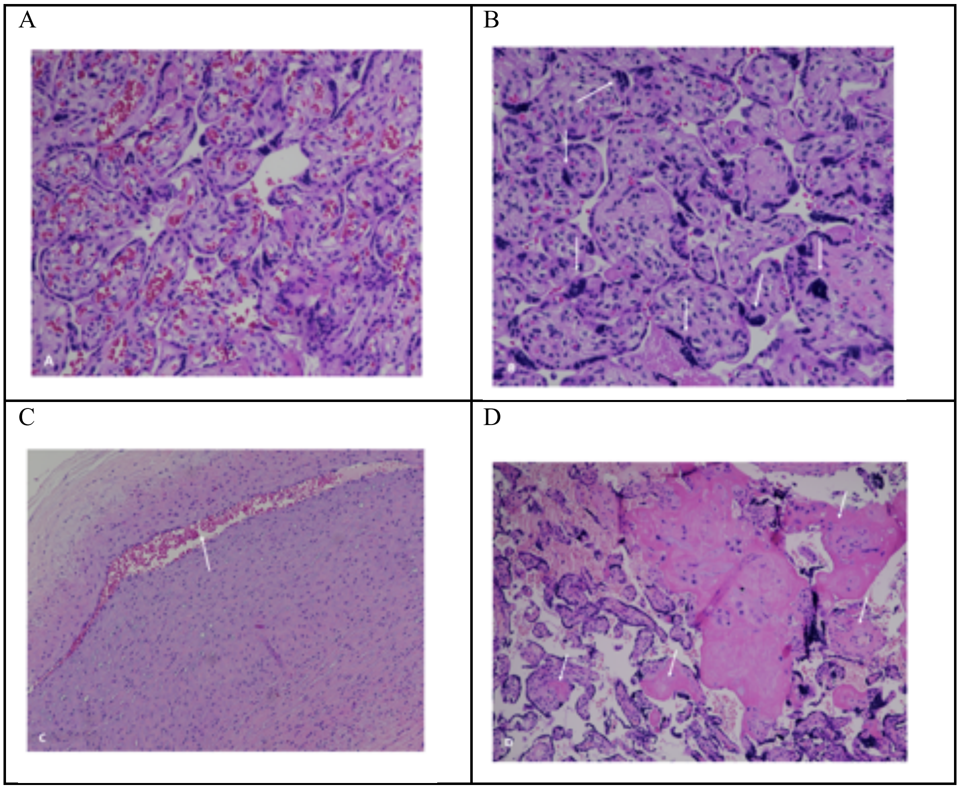

Supplementary Figure S5. Placental Samples Examination. hematoxylin-eosin, original magnification X10

A- Normal histologic appearance of placental villi (from a vaccinated patient)

B- Maternal vascular malperfusion lesion- increased syncytial knots (arrows) (from past covid 19 infected parturient)

C- Fetal vascular malperfusion lesion- chorionic vessel with intramural fibrin deposition. Arrow towards vessel lumen (from a vaccinated patient)

D- Maternal vascular malperfusion lesion- increased intervillous fibrin deposition (arrows) (from past covid 19 infected parturient)

### Supplementary Tables

Supplementary Table S1

**Placental Samples Examination**

Placental pathology examinations were performed on randomly taken placental tissue samples of 1X1 $cm^3$ thickness, that were fixed in formalin and embedded in paraffin blocks for microscopic assessment. All examinations were done by a single pathologist (author L.S.), who was blinded to the study groups the samples were taken from. Placental lesions were divided into 3 main groups: 1. Maternal vascular malperfusion (MVM) lesions that included: decidual vasculopathy, increased syncytial knots, villous agglutination, increased intervillous fibrin deposition, and villous infarcts; 2. Fetal vascular malperfusion (FVM) lesions that included large vessel thrombosis or fetal thrombotic vasculopathy, hypovascular, fibrotic and avascular villi; 3. Chronic villitis that was defined as villitis of unknown etiology, plasma cell deciduitis, or chronic intervillositis;

Fourteen placental tissue samples were microscopically examined. The rate of malperfusion lesions was similar in the examined placental tissue

| Placental lesions | Control<br>n=5 | Past Covid -19<br>n=3 | Vaccinated<br>n=6 |
|---|---|---|---|
| MVM | 1 | 2 | 2 |
| FVM | 2 | 0 | 2 |
| Chronic villitis | 0 | 0 | 1 |

**Supplementary Table S2**. Statistical analysis of maternal and fetal serological response to vaccination. Analysis of the data presented in Figure 2D

| Ab type | Comparison | Statistical test | P value |
|---|---|---|---|
| **IgG S1** | Maternal Ab concentrations among groups | Kruskall–Wallis one-way ANOVA, followed by Dunn's All-Pairwise Comparisons Test. | < 0.0001 |
| | Fetal Ab concentrations among groups | Kruskall–Wallis one-way ANOVA, followed by Dunn's All-Pairwise Comparisons Test. | < 0.0001 |
| | Maternal vs. fetal Ab concentrations within the Control group | Paired t-test | 0.0034 |
| | Maternal vs. fetal Ab concentrations within the 1st Dose group | Paired t-test | 0.0050 |
| | Maternal vs. fetal Ab concentrations within the 2nd Dose group | Paired t-test | 0.0071 |
| | Maternal vs. fetal Ab concentrations within the Fully-vaccinated group | Paired t-test | 0.0085 |
| **IgG S2** | Maternal Ab concentrations among groups | Kruskall–Wallis one-way ANOVA, followed by Dunn's All-Pairwise Comparisons Test. | < 0.0001 |
| | Fetal Ab concentrations among groups | Kruskall–Wallis one-way ANOVA, followed by Dunn's All-Pairwise Comparisons Test. | < 0.0001 |
| | Maternal vs. fetal Ab concentrations within the Control group | Paired t-test | 0.4190 |
| | Maternal vs. fetal Ab concentrations within the 1st Dose group | Paired t-test | 0.0203 |
| | Maternal vs. fetal Ab concentrations within the 2nd Dose group | Paired t-test | 0.0930 |
| | Maternal vs. fetal Ab concentrations within the Fully-vaccinated group | Paired t-test | 0.0197 |

| | | | |
|---|---|---|---|
| **IgG RBD** | Maternal Ab concentrations among groups | Kruskall–Wallis one-way ANOVA, followed by Dunn's All-Pairwise Comparisons Test. | < 0.0001 |
| | Fetal Ab concentrations among groups | Kruskall–Wallis one-way ANOVA, followed by Dunn's All-Pairwise Comparisons Test. | < 0.0001 |
| | Maternal vs. fetal Ab concentrations within the Control group | Paired t-test | 0.0084 |
| | Maternal vs. fetal Ab concentrations within the 1st Dose group | Paired t-test | 0.0052 |
| | Maternal vs. fetal Ab concentrations within the 2nd Dose group | Paired t-test | 0.0069 |
| | Maternal vs. fetal Ab concentrations within the Fully-vaccinated group | Paired t-test | 0.0339 |
| **IgG N** | Maternal Ab concentrations among groups | Kruskall–Wallis one-way ANOVA, followed by Dunn's All-Pairwise Comparisons Test. | 0.3016 |
| | Fetal Ab concentrations among groups | Kruskall–Wallis one-way ANOVA, followed by Dunn's All-Pairwise Comparisons Test. | 0.3869 |
| | Maternal vs. fetal Ab concentrations within the Control group | Paired t-test | 0.0142 |
| | Maternal vs. fetal Ab concentrations within the 1st Dose group | Paired t-test | 0.0562 |
| | Maternal vs. fetal Ab concentrations within the 2nd Dose group | Paired t-test | 0.2313 |
| | Maternal vs. fetal Ab concentrations within the Fully-vaccinated group | Paired t-test | 0.08903 |
| **Ab type** | **Comparison** | **Statistical test** | **P value** |
| **IgM S1** | Maternal Ab concentrations among groups | Kruskall–Wallis one-way ANOVA, followed by Dunn's All-Pairwise Comparisons Test. | < 0.0001 |
| | Fetal Ab concentrations among groups | Kruskall–Wallis one-way ANOVA, followed by Dunn's All-Pairwise Comparisons Test. | 0.1176 |

| | | | |
|---|---|---|---|
| | Maternal vs. fetal Ab concentrations within the Control group | Paired t-test | < 0.0001 |
| | Maternal vs. fetal Ab concentrations within the 1st Dose group | Paired t-test | < 0.0001 |
| | Maternal vs. fetal Ab concentrations within the 2nd Dose group | Paired t-test | < 0.0001 |
| | Maternal vs. fetal Ab concentrations within the Fully-vaccinated group | Paired t-test | < 0.0001 |
| **IgM S2** | Maternal Ab concentrations among groups | Kruskall–Wallis one-way ANOVA, followed by Dunn's All-Pairwise Comparisons Test. | 0.0417 |
| | Fetal Ab concentrations among groups | Kruskall–Wallis one-way ANOVA, followed by Dunn's All-Pairwise Comparisons Test. | 0.7675 |
| | Maternal vs. fetal Ab concentrations within the Control group | Paired t-test | < 0.0001 |
| | Maternal vs. fetal Ab concentrations within the 1st Dose group | Paired t-test | < 0.0001 |
| | Maternal vs. fetal Ab concentrations within the 2nd Dose group | Paired t-test | < 0.0001 |
| | Maternal vs. fetal Ab concentrations within the Fully-vaccinated group | Paired t-test | < 0.0001 |
| **IgM RBD** | Maternal Ab concentrations among groups | Kruskall–Wallis one-way ANOVA, followed by Dunn's All-Pairwise Comparisons Test. | 0.0067 |
| | Fetal Ab concentrations among groups | Kruskall–Wallis one-way ANOVA, followed by Dunn's All-Pairwise Comparisons Test. | 0.0560 |
| | Maternal vs. fetal Ab concentrations within the Control group | Paired t-test | < 0.0001 |
| | Maternal vs. fetal Ab concentrations within the 1st Dose group | Paired t-test | < 0.0001 |

| | | | |
|---|---|---|---|
| | Maternal vs. fetal Ab concentrations within the 2nd Dose group | Paired t-test | < 0.0001 |
| | Maternal vs. fetal Ab concentrations within the Fully-vaccinated group | Paired t-test | < 0.0001 |
| **IgM N** | Maternal Ab concentrations among groups | Kruskall–Wallis one-way ANOVA, followed by Dunn's All-Pairwise Comparisons Test. | 0.9558 |
| | Fetal Ab concentrations among groups | Kruskall–Wallis one-way ANOVA, followed by Dunn's All-Pairwise Comparisons Test. | 0.1804 |
| | Maternal vs. fetal Ab concentrations within the Control group | Paired t-test | < 0.0001 |
| | Maternal vs. fetal Ab concentrations within the 1st Dose group | Paired t-test | < 0.0001 |
| | Maternal vs. fetal Ab concentrations within the 2nd Dose group | Paired t-test | < 0.0001 |
| | Maternal vs. fetal Ab concentrations within the Fully-vaccinated group | Paired t-test | < 0.0001 |
